# Tempo-spatial dynamics of COVID-19 in Germany – A phase model based on a pandemic severity indicator

**DOI:** 10.1101/2023.02.17.23286084

**Authors:** Martin Stabler, Andreas Kuebart

**Affiliations:** Leibniz Institute for Research on Society and Space; Free University of Berlin; Brandenburg Technical University

## Abstract

While pandemic waves are often analyzed on the national scale, they typically are not distributed evenly in space. This working paper employs a novel approach to analyze the tempo-spatial dynamics of the COVID-19 pandemic for the case of Germany. First, we base the analysis not just on the incidence of cases or mortality but employ a composite indicator of pandemic severity to gain a more robust understanding of the temporal dynamics of the pandemic. Second, we subdivide the pandemic during the years 0f 2020 and 2021 into fifteen phases, each with a coherent trend of pandemic severity. Third, we analyze the spatial patterns predominating in each phase. The resulting tempo-spatial phase model is used to identify explanatory factors for the tempo-spatial patterns identified in the analysis.

**Disclosures:** This work was supported by the Deutsche Forschungsgemeinschaft (DFG, German Research Foundation) with a research grant for the project Socio-spatial diffusion of COVID-19 in Germany. - 492338717

## Introduction

The COVID-19 pandemic, like any other pandemic, follows a pattern of acceleration and deceleration concerning infection rates and incidence values among populations respectively. Over the course of a pandemic, this creates a temporal pattern in form of logistic curves often described as wave(s). These waves consist of (fast) ascents, (up to several) peaks, and declines that indicate several stages or phases of a pandemic. While pandemic waves are often analyzed on the national scale, they typically are not distributed evenly within territories (Cliff et al. 2009; Śleszyński 2021; Teller 2021; Boterman 2022; Keeler & Emch 2018). In contrast, the spatial patterns of infections and pandemic severity vary over time, which is why a tempo-spatial perspective is necessary to understand the spread of infectious diseases (Ghosh&Cartone 2021). In the case of COVID-19, several studies have found COVID-19 infections to be clustered within countries (Scarpone et al. 2020, Murgante et al. 2020, Rodríguez-Pose & Burlina 2021), and even within cities (Slijander et al. 2021). This paper aims to trace the tempo-spatial patterns of the COVID-19 pandemic in Germany throughout the years 2020 and 2021. COVID-19 was first detected in Germany in January 2021, with four pandemic waves occurring in the first two years of the pandemic.

To explain the tempo-spatial dynamics of the COVID-19 pandemic in Germany, we proceeded in four analytical steps: First, we develop a composite index of ‘pandemic severity’, which integrates the three sub-indicators of COVID-19 case incidence, the incidence of death due to COVID-19, and the incidence of COVID-19 patients in intensive care. Second, a phase model based on the time series of a pandemic severity indicator is developed based on a change point analysis. Each of the fifteen stages in the model is coherent in terms of the trends of pandemic dynamics (e.g., rising, decreasing, stable). Third, the spatial pattern during each phase is analyzed by considering global and local spatial autocorrelation. To analyze the tempo-spatial variation of pandemic severity we thus opted to analyze the spatial patterns of pandemic severity during the whole phases instead of relying on snapshots on specific dates. The hotspot and cold spot regions identified for each phase are then related to the drivers of pandemic severity. To do so, we combined time-series analysis on the national scale with hotspot analysis on the regional scale (NUTS 3). The paper is structured as follows: The following section describes how we proceeded and highlights methodological reasoning for the pandemic severity index, the phase model, and the spatial analysis. The next section presents the resulting time series and profiles each of the fifteen individual phases. The final section discusses the results and concludes.

## Data & Methods

### Data collection

All datasets for the pandemic severity index were sourced via Corona Daten Plattform^1^., Data on COVID-19 cases and deaths originate from the federal Robert Koch Institute, and data on patients with COVID-19 in intensive care were used as reported by DIVI. All indicators were available on the NUTS3 level, which represents 400 counties and cities (Kreise and Kreisfreie Städte). For the initial change point analysis the dataset was aggregated to the national scale. Additional datasets were used to further describe the phases but are not part of the composite indicator. These include data on outbreaks, and data on the mobility of the population during the pandemic, both provided by RKI (see Schlosser et al. 2020), as well as the stringency index for Germany provided by the Oxford COVID-19 Government Response Tracker (Hale et al. 2020). Data on the vaccination campaign was provided by RKI, while data on dominating strains of SARS-CoV-2 was sourced from Hodgecroft (2021).

### A composite index for pandemic severity

Studies focusing on the tempo-spatial aspects of the COVID-19 pandemic rely on the case incidence of patients tested positive for COVID-19 almost exclusively as an indicator (Nazia et al. 2022). Recently, it has been suggested to combine several indicators when researching the spatio-temporal analysis of COVID-19 (Pagel & Yates 2021; Rohleder & Bozorgmehr 2022). Therefore, we decided to do so to add robustness, especially since the testing regime changed throughout the pandemic and potentially also varied regionally. For example, during the first wave of COVID-19 in Germany (March through May 2020), fairly limited capacities of PCR testing were available, while later PCR testing and antigen testing were widely available.

The pandemic severity index consists of three indicators: the incidence of patients tested positive for COVID-19, the incidence of patients with COVID-19 in intensive care, and the incidence of registered deaths due to COVID-19 (see table 1). The process of designing a composite indicator includes the scaling, weighting, and aggregating of the initial datasets (Liborio et al. 2021). The pandemic severity index was calculated as follows: First, the rolling 14-day mean of each sub-indicators was calculated by using the zoo package in R (Zeileis & Grothendieck 2005) before the sub-indicators were normalized by using z-scores. Second, the indicators were scaled by a minimum-maximum normalization to values between 0 and 1 (Wickham & Seidel 2022). Third, the indicators were aggregated by using the arithmetic mean, so that all three sub-indicators have the same weight. Subsequently, the composite index was rounded to four decimal places. This approach represents the common procedure to calculate composite indicators (Dialga & Giang 2017). The resulting pandemic severity index indicates the pandemic burden during a given day in each county through a value between 0 and 1 (in practice between 0 and 0.9326). It was calculated for each day between 2020-03-01 and 2021-12-31 for each German county / NUTS3 region, resulting in 268,400 individual values. To establish a phase model on the national scale, it was aggregated for each day.

**Table 1:** Elements of the pandemic severity composite indicator

| Indicator | Time range | Spatial resolution |
| --- | --- | --- |
| Patients tested positive for COVID-19 | 2020-02-01 -<br>2021-12-31 | NUTS 3 |
| Deaths of patients tested positive for COVID-19 | 2020-02-01 -<br>2021-12-31 | NUTS 3 |
| Patients tested positive for COVID-19 in intensive care | 2020-03-04 -<br>2021-12-31 | NUTS 3 |

### A phase model based on change points of pandemic severity

A comprehensible method for delineating pandemic stages is rarely found in the literature on COVID-19. Most existing phase models (Ghosh & Cartone, 2020; Benita & Gasca-Sanchez 2021, Li et al. 2021, Zawbaa et al. 2022) define the beginning of each stage based on individual indicators, such as incidence rates, mortality rates or the implementation of counter measures like lockdowns and social distancing. Typical types of phases include ‘beginning’, ‘outbreak’, ‘recession’ and ‘plateau’ (Li et al. 2021). Schilling et al. (2022) use a multivariant approach by combining several variables for their phase model for Germany, however their method of delineating phases remains unclear. Küchenhoff et al. (2021) calculate change points in the early course of the pandemic from March to May 2020 using a back-projection for estimated daily infections in Germany and Bavaria. Their retrospective exploratory analysis identified five phases for Germany and six phases for Bavaria respectively, within the first wave.

Our phase model of the COVID-19 pandemic in Germany was developed by performing a two-step change point analysis based on a time series of the pandemic severity index on the national scale. The change point analysis was performed by using the binary segment approach (Scott & Knott 1974) on mean and variance of the lagged difference of the pandemic severity indicator. The first step established rough phases, which then served as a heuristic to develop a fine-grained model. Here. a minimum segment length of fourteen days was used with the binary segment approach. The change points were calculated using the change point package in R (Killick & Eckley 2014). The resulting phase model consists of fifteen individual phases that range from 19 to 116 days in length.

For each phase, several explanatory indicators were calculated, including the average stringency index of policy responses against the pandemic, average mobility patterns, number and settings of prominent outbreaks, progress of the vaccination campaign, and the distribution of SARS-CoV-2 clades in Germany during the respective phase.

### Local and global autocorrelation

Subsequent to the temporal change point analysis, a spatial analysis was performed in three steps: First, the average pandemic severity in each NUTS3 region was calculated for each phase and mapped. Second, the global spatial autocorrelation of pandemic severity was calculated for each day in the study period in form of Moran’s I metric (Moran 1950). Moran’s I and the related Moran’s I test statistic were calculated by using the sfdep package in R (Parry 2022). Third, each of the fifteen phases were analyzed in terms of their individual spatial patterns: Local indicators of spatial association (LISA) were calculated for each phase using localized Moran’s I (Anselin 1995, Sokal et al. 1998) using the sfdep package in R (Parry 2022). The LISA analysis was performed based on a contiguity network without weights. LISA analysis has been used before to identify significant clusters of pandemic outbreaks (e.g., Scarpone et al. 2020, Siljander et al. 2022). However, in this study the LISA analysis is based on a composite indicator of pandemic severity instead of case incidence or mortality as single indicators. A LISA analysis groups the spatial units (here counties / NUTS3 regions) relative to their neighbors in local clusters of high values (high–high) or low values (low–low), and also identifies spatial outliers with (high–low) or (low–high) values. For each spatial unit, the p-value was determined through 499 simulations. Only results with a p-value below 0.05 were considered. Since the geography of the COVID-19 pandemic has been found to be uneven in space (Scarpone et al. 2020, Rodríguez-Pose & Burlina 2021), the pandemic severity index can also help to reveal the complex tempo-spatial patterns of how the pandemic unfolded better compared to case numbers. Visualizations were created in R using the ggplot2 (Wickham 2016), and sf (Pebesma 2018) packages.

### A Phase model of the COVID-19 pandemic in Germany

The fifteen phases of our model (table 1, chart 1) are based on trend coherence and thus differ in length. On average, each phase lasts about 47 days, although the longest phase is more than twice as long (summer plateau 2020: 116 days) and the shortest lasted only 19 days (surge of the first wave). Since the phases are based on a change point analysis using the lagged daily difference of the pandemic severity index, pandemic waves consist at least of two phases (increase and decrease). However, more complex waves can consist of several intermediate phases of acceleration and deceleration, as for example the second COVID-19 wave in Germany consist of four individual phases. Further, the periods between the phases are also considered as individual phases. This relatively fine-grained phase model enables more specific insights into tempo-spatial patterns, which indeed vary substantially between the different phases within one wave.

**Table 2:** Overview over the fifteen phases of the phase model

| Sub-phase | Duration | Length in days |
| --- | --- | --- |
| A Prelude first wave 2020 | 2020-02-01 - 2020-03-19 | 48 |
| B Surge first wave 2020 | 2020-03-20 - 2020-04-07 | 19 |
| C Decline first wave 2020 | 2020-04-08 - 2020-05-25 | 48 |
| D Plateau summer 2020 | 2020-05-26 - 2020-09-18 | 116 |
| E Entry Surge winter wave 2020/21 | 2020-09-19 - 2020-10-10 | 22 |
| F Surge winter wave 2020/21 | 2020-10-12 - 2020-11-13 | 33 |
| G Further surge winter wave 2020/21 | 2020-11-14 - 2020-12-23 | 40 |
| H Decline winter wave 2020/21 | 2020-12-24 - 2021-03-02 | 69 |
| I Surge easter wave 2021 | 2021-03-03 - 2021-04-28 | 57 |
| J Decline easter wave 2021 | 2021-04-29 - 2021-06-16 | 49 |
| K Bottom summer 2021 | 2021-06-17 - 2021-07-17 | 31 |
| L Entry surge summer 2021 | 2021-07-18 - 2021-08-10 | 24 |
| M Entry delta wave 2021 | 2021-08-11 - 2021-10-24 | 75 |
| N Surge delta wave 2021 | 2021-10-25 - 2021-12-02 | 39 |
| O Decline delta wave 2021 | 2021-12-03 - 2021-12-30 | 28 |

**Figure 1:**
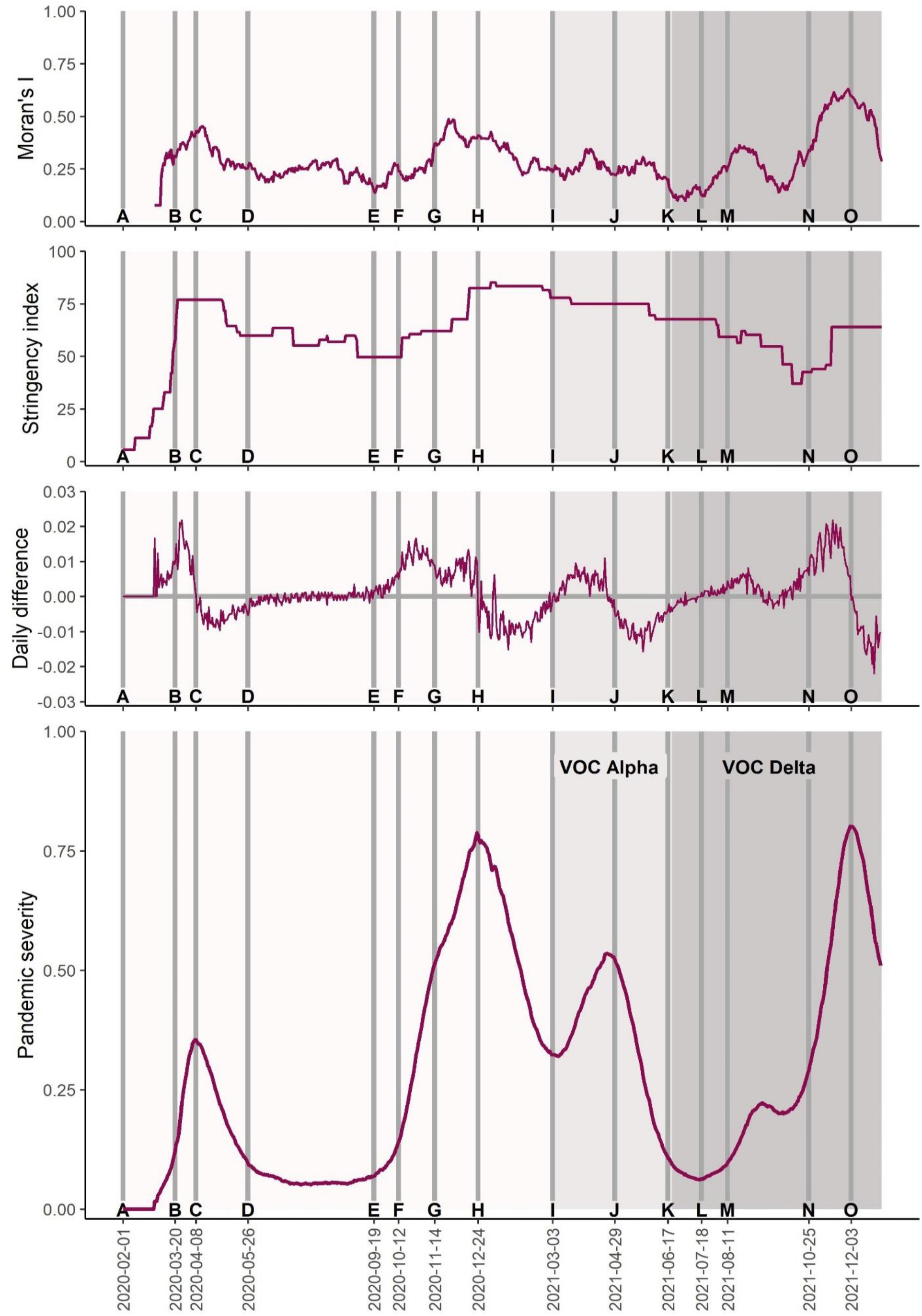
Temporal dynamics of COVID-19 in Germany during 2020 and 2021. The figure shows the trajectories of the pandemic severity composite indicator (bottom), the daily difference, the stringency of non-pharmaceutical interventions and the spatial autocorrelation (top). In each chart, the fifteen phases of the phase model are highlighted together with the dominant strain of SARS-CoV-2

### A Prelude first wave 2020-01-27 - 2020-03-19

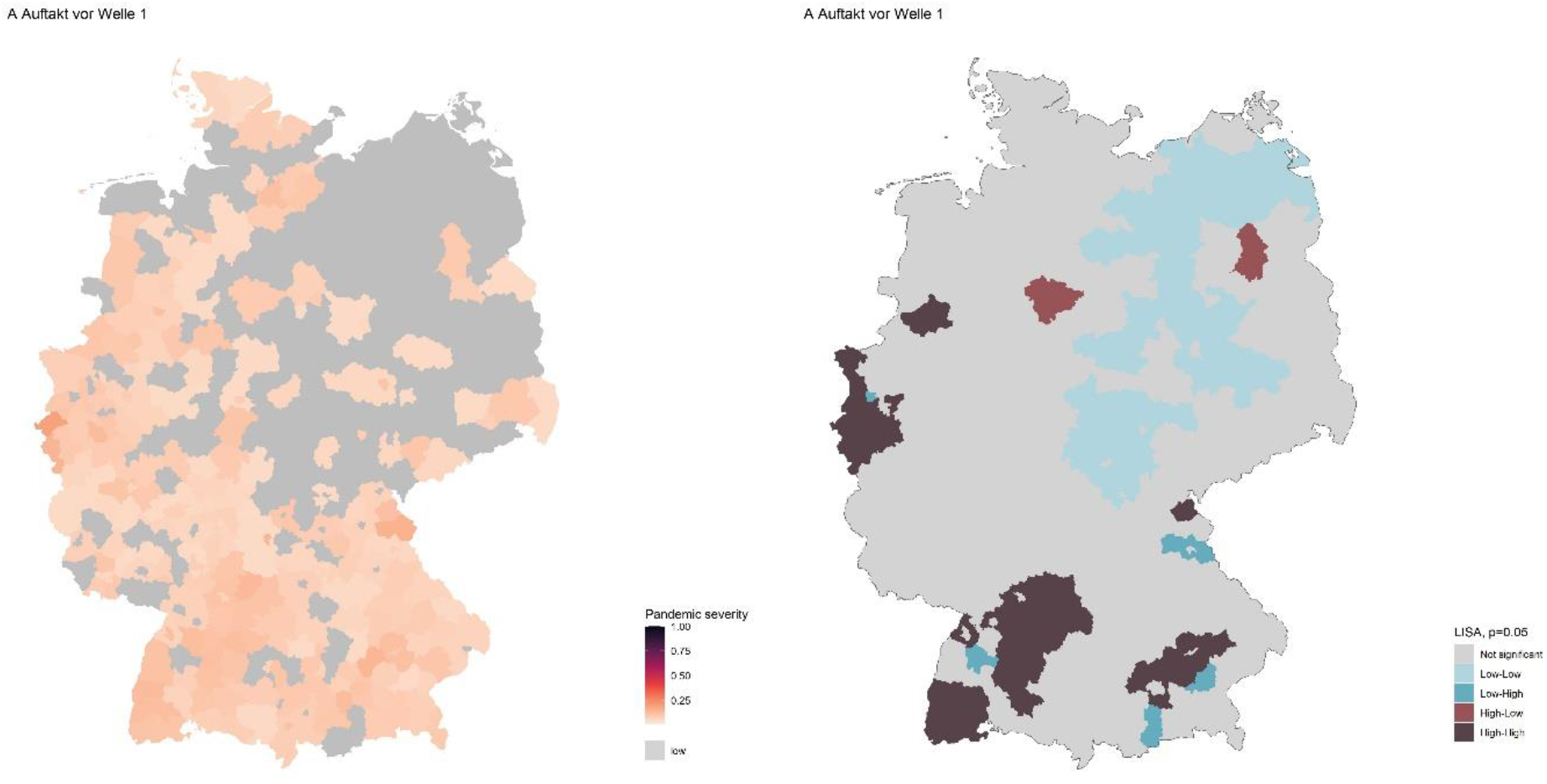

The (then) novel SARS-CoV-2 was first detected in Germany in late January in relation to a cluster of infections in the Munich region. The following 52 days, where characterized by a low level of pandemic severity that started to increase rapidly at the beginning of March, although the data on hospitalizations limits the effectiveness of the pandemic severity indicator until early March. In February, the social life was still relatively unhampered. For example, the 2020 carnival season was held without major precautions. However, the first non-pharmaceutical interventions, such as precautionary quarantines for travelers entering form China were already in place. Around mid-March, strict counter measures including border closures, closure of various public venues and shortly after strict social distancing were implemented. The early dynamics of pandemic diffusion were triggered by regional outbreaks on the one hand and infections of returnee infections after vacations on the other hand (Kuebart & Stabler 2020A). Private and public festivities played an important role for super spreading events causing regional outbreaks. Notable examples include a carnival event in Heinsberg county in North-Rhine Westphalia, a party at a nightclub in Berlin, and a beer festival in Tirschenreuth county in Bavaria. Tourist returnees from ski resorts in Austria and Italy were a major driver for rising pandemic severity in large parts of southern and western Germany. The modest degree of spatial autocorrelation during this phase implies a comprehensive introduction of the new pathogens during the first phase, limiting the relevance of regional outbreaks for the further advance of the pandemic.

### B Surge first wave 2020-03-20 - 2020-04-07

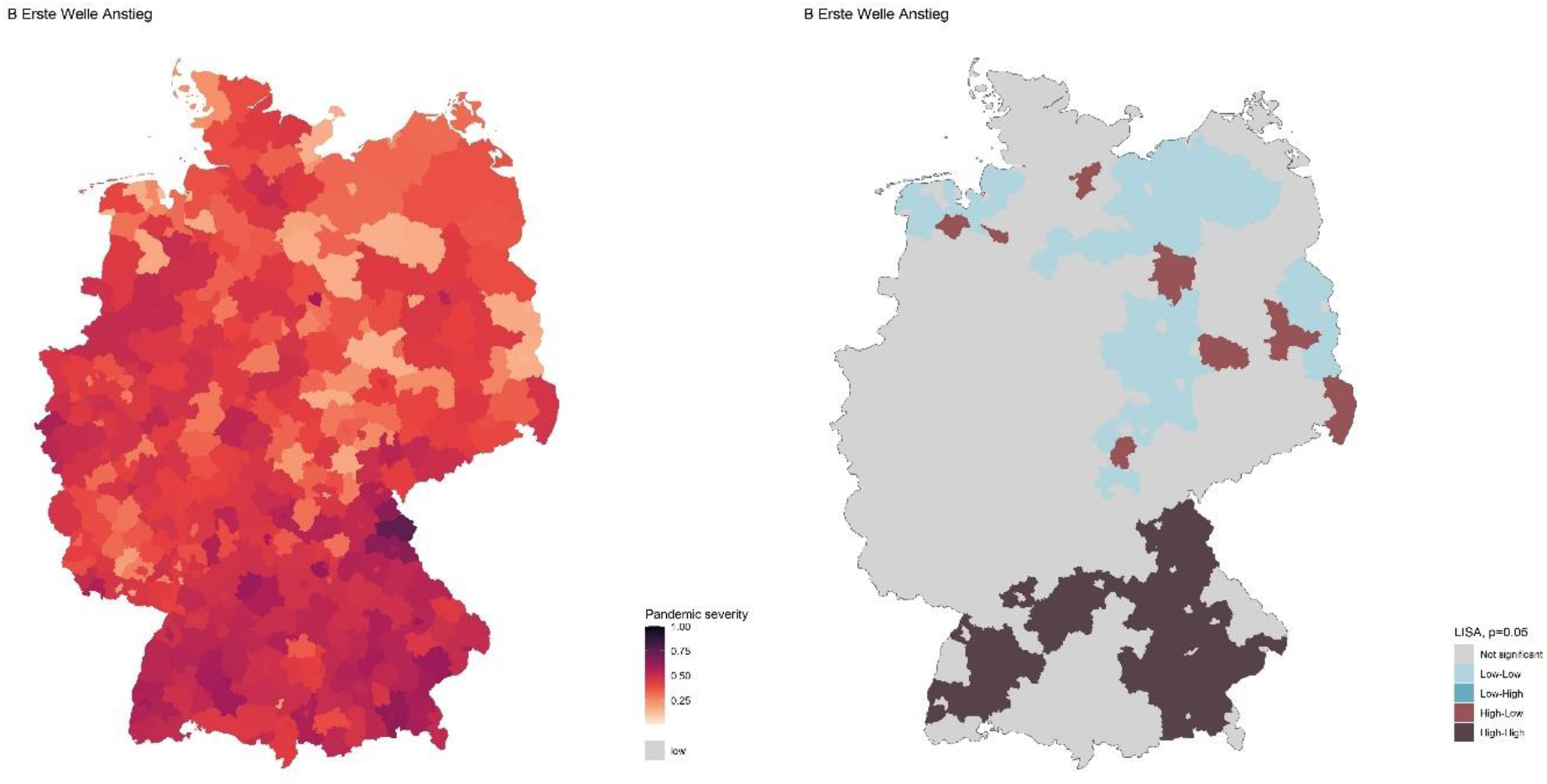

The length of the second stage of the pandemic is 19 days. The surge of the first wave 2020 started on the 20^th^ of March and lasts until the 7^th^ of April 2020. This phase is dominated by the first surge of SARS-CoV-2. The surge of the first wave has been the shortest phase in this phase-model since the duration of rapid exponential growth was only 19 days. This time was characterized by a period of exponential growth of pandemic severity, resulting in a relatively rapid spread of the virus, so that each German region was hit. However, the overall level of pandemic severity was still low compared to subsequent waves. After the introduction of strict non-pharmaceutical interventions (the stringency index rose from 25 on 2020-03-08 to 76.85 on 2020-03-22) the exponential growth flattened towards the end of this phase. Reduced contacts and a virtual shutdown of social life result in a phase with the lowest average mobility values throughout the pandemic.

Spatially, the distribution of the severity index shows a relatively high spatial clustering. Especially in southern Germany spatial clustering is prominent with a pronounced hotspot around the county of Tirschenreuth. The peak of the first COVID-19 wave marks the end of this stage. In contrast, much of northern Germany is dominated by cold spots as described by Scarpone et al. (2020).

### C Decline first wave 2020-05-26 - 2020-09-18

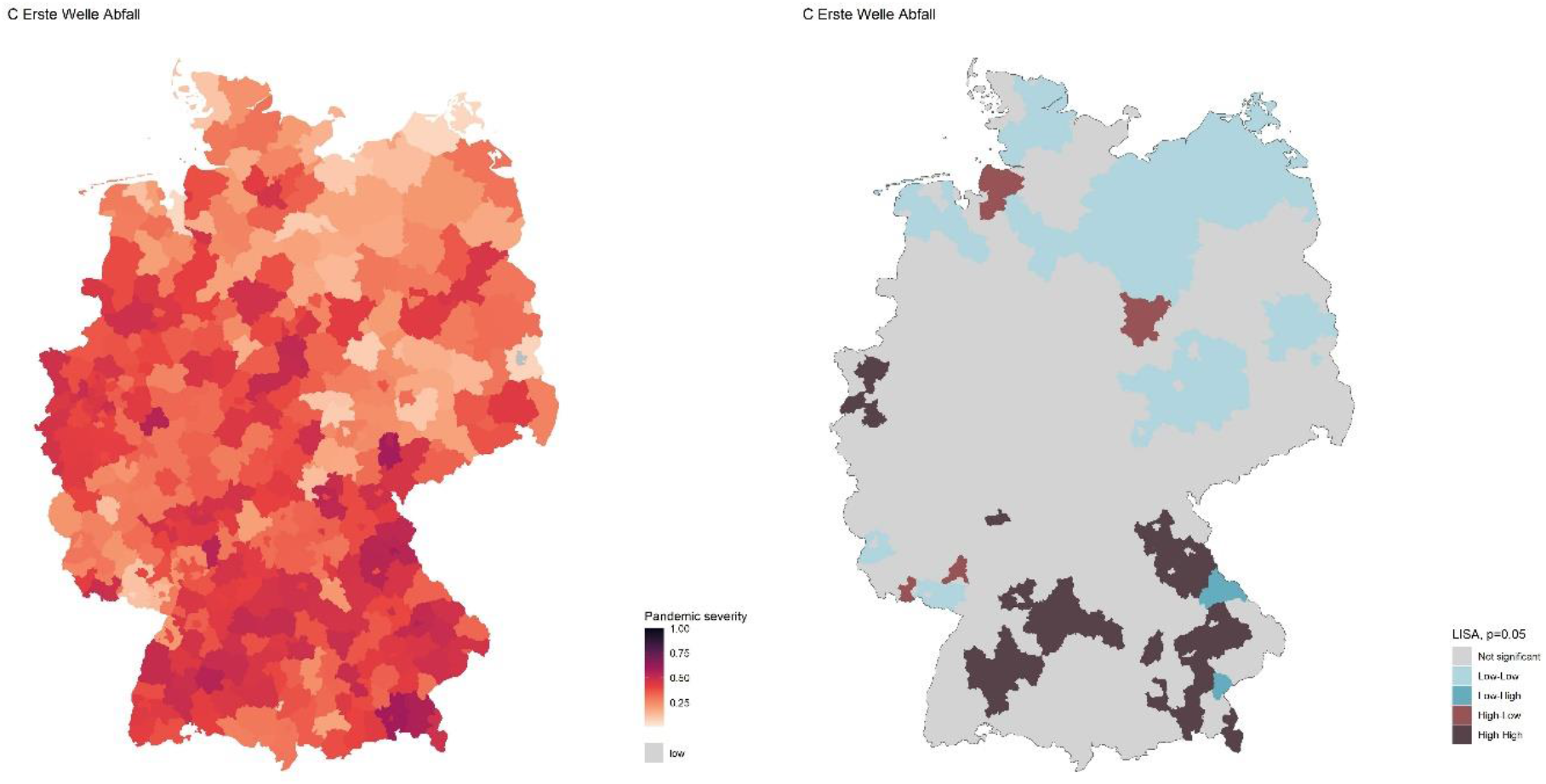

The third stage delineates the decline of the first wave. It lasted 48 days in the spring of 2020. The decline reflects the effectiveness of the containment measures introduced during the surge of the first wave in mid-March 2020. The drastic reduction of social contacts and a shutdown of social life limited the spread of the disease with a time lag of about two weeks. The stringency of non-pharmaceutical interventions was only lowered towards the end of this phase in early May. Only then the number of average contacts also started to slowly increase but remained on a level about a third below the average before the pandemic.

Therefore, the decline of the first wave was not caused by saturation effects, but rather the cutting of infection networks and especially the inhibition of super-spreading events. However, outbreaks in closed environments such as elderly care facilities dominated this phase, with about 20% of reported cases belonging to this category, which is the highest value over all fifteen phases. The raw case fatality rate was very high in both this and the preceding phase. At above 5% the third phase has the highest value for all phases. This might be caused by less frequent testing and thus lower case-detection-rates on the one hand, and less effective treatments compared to later stages of the pandemic on the other hand.

The geography of hot spot regions remains largely in place compared to the previous phase. The regions in southern Germany that have been hit hardest still had above average levels of pandemic severity while individual regions in western and northern Germany saw a relative spike, possibly due to outbreaks in closed environments. The cold spots in northern Germany prevail mostly, though.

### D Plateau summer 2020-05-26 - 2020-09-18

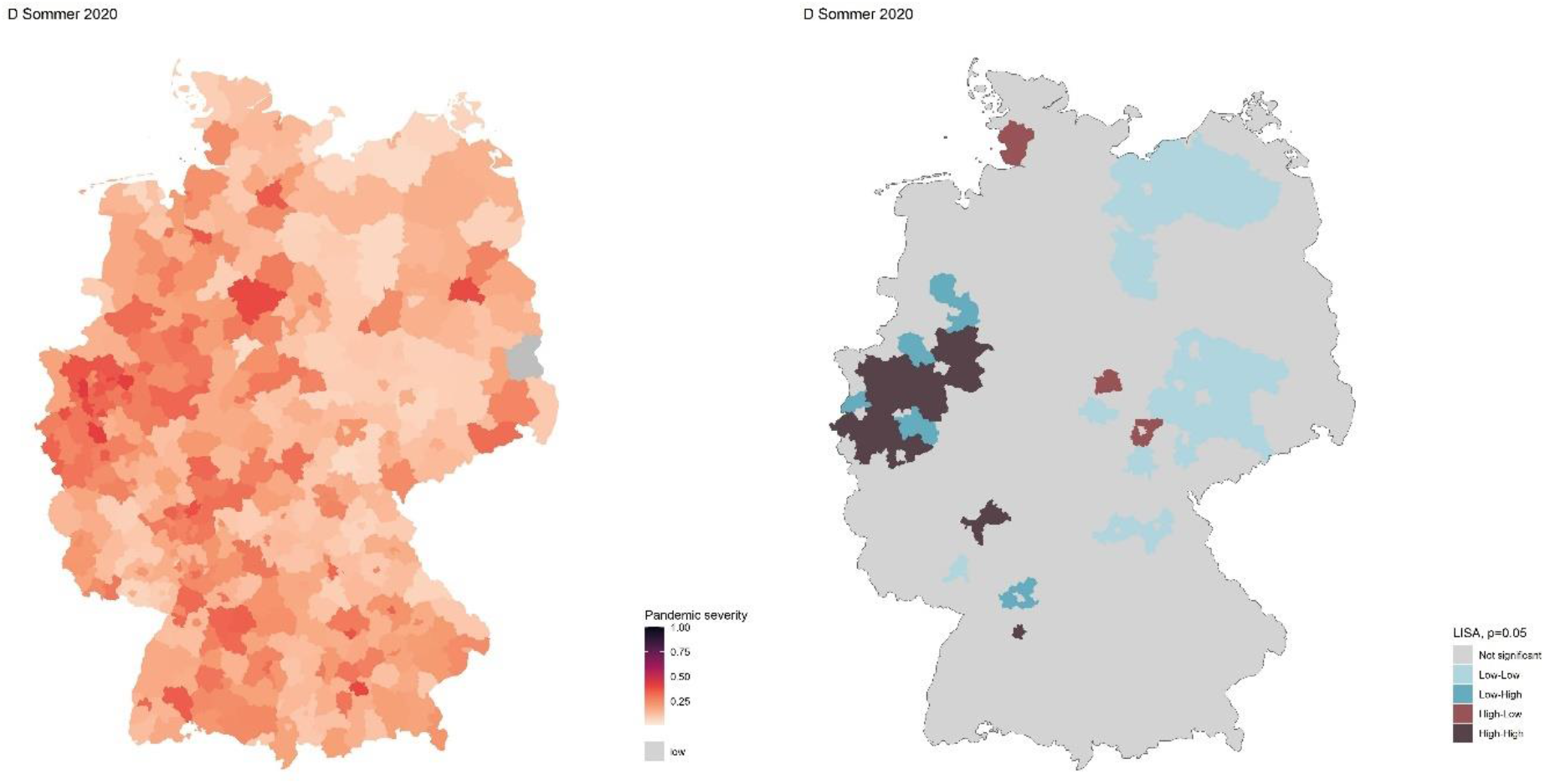

With 116 days, the fourth stage is by far the longest of all fifteen stages. It is defined by the summer plateau after the first wave during the summer months in 2020. The pandemic severity index hovers at relatively low levels and only begins to climb in early September after the summer holidays have ended in most German regions. While the incidence level was extremely low during this period, the numbers of intensive care patients and deaths due to COVID-19 did not decrease that heavily. This still implies shortcomings in the case detection regime during this period. Contacts and individual mobility increased throughout the phase but did not reach pre-pandemic values, since many measures did stay in place.

However, the increase in social contacts also enabled more super-spreading events, resulting in a relatively high share of cases related to outbreaks in crowded social settings. This is reflected in the spatial structure of pandemic severity during the fourth phase, which is characterized through a number of spatially scattered outbreaks. Among them a meat processing factory that accounts for more than 2000 infections in Gütersloh county. Further, several cities and counties in North-Rhine Westphalia emerge as hotspots, possibly due to imported cases during the summer vacation season (Frank et al. 2021). Overall, the spatial autocorrelation of pandemic severity is lower compared to the previous phases.

### E Entry winter wave 2020-09-19 - 2020-10-10

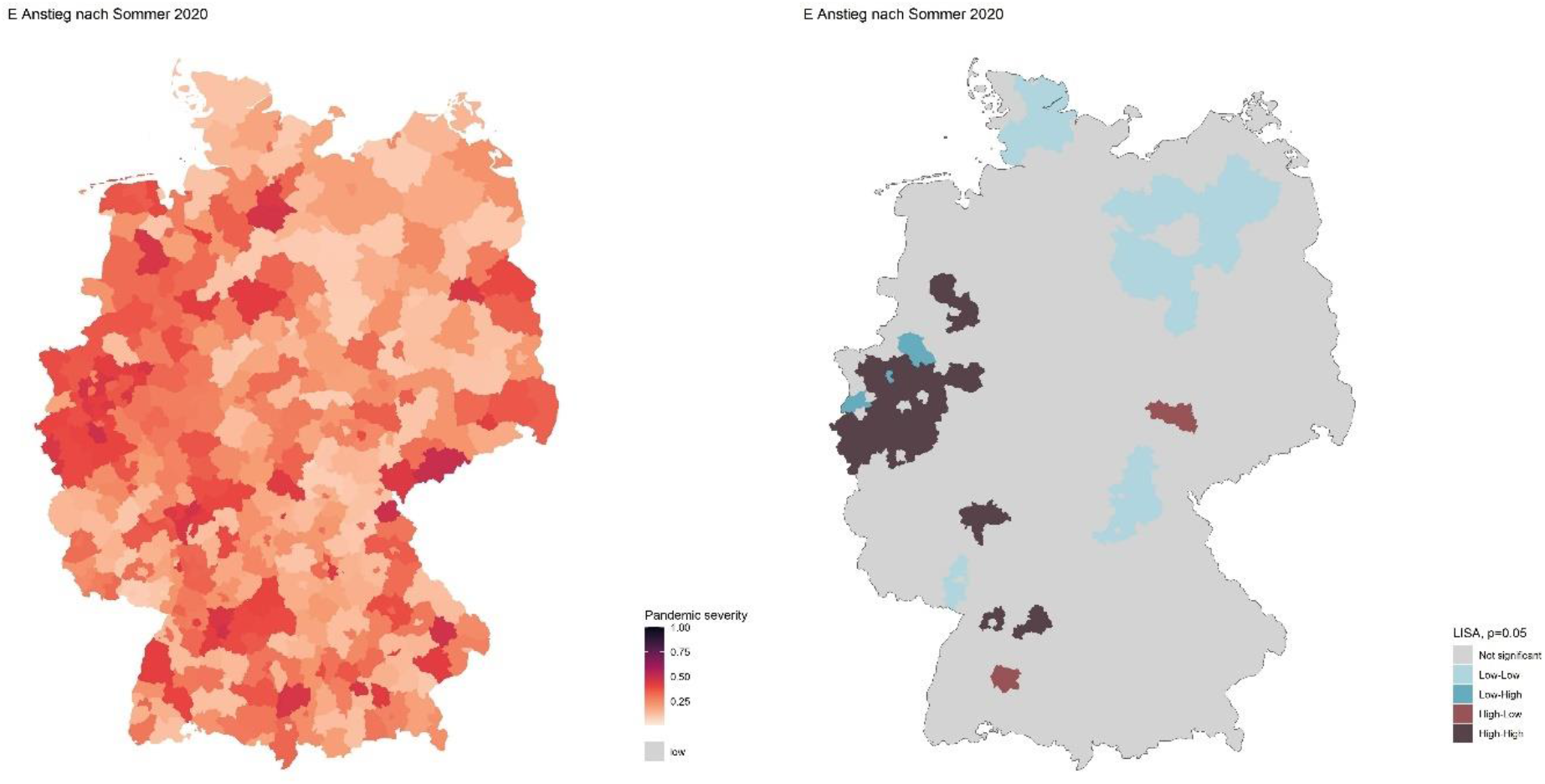

The fifth stage describes the transition between the summer plateau and the surge in autumn and winter 2020. Thus, these three weeks in September and October set the conditions for the second wave. During the summer plateau in 2020, the strictness of pandemic response policies had declined substantially, so that during this phase the stringency index has the lowest values among of all phases. A relatively lax atmosphere is also evident in a high social contact index, which is during this phase on average about twice as high as during the surge of the first wave. Further, this phase is characterized by low spatial autocorrelation. The hot spots and cold spots mostly persist compared to the previous phase.

### F Winter surge 2020-10-12 - 2020-11-13

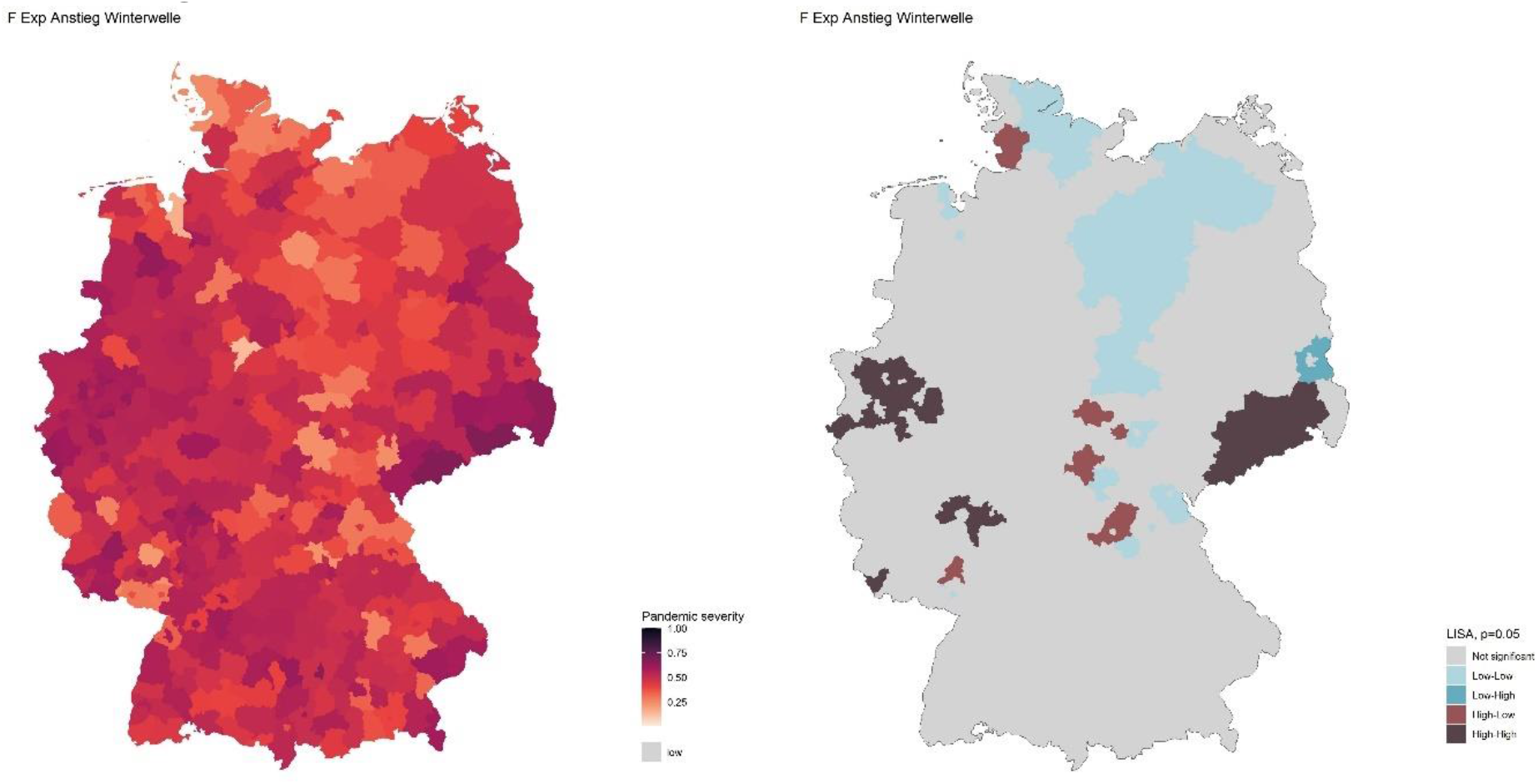

After the modest increase during the previous phase, the winter surge accelerated considerably., so that the severity index increases at an unprecedented rate. The phase between mid-October and mid-November 2020 is characterized by a similarly rapid increase in pandemic severity as during phase B. However, this phase lasted about two weeks longer than the surge of the first wave was not followed by an immediate decline. Unlike later surges, the rapid increase in pandemic severity during this phase was not caused by the introduction of a new variant of COVID-19. In contrast, there were several SARS-CoV-2 variants circulating in Germany at that time, especially from the clades B.1.177 and B.1.^2^

The rapid increase in pandemic severity induced a (re)-introduction of severe non-pharmaceutical measures and thus an increase in the stringency index right at the beginning of this phase. While a “stay at home” policy was announced by chancellor Merkel on October 17^th^, the contact index does not show an immediate reduction of average social contacts, indicating a lack in compliancy. Instead, only a trend of steady reduction of social contacts is triggered that continues until March 2021.

Despite the surge in pandemic severity, the spatial autocorrelation remains on a low level. This implies that the surge is not triggered by specific clusters like the previous surge (phase B), but rather the severity index increased in wide parts of Germany. However, the cold spots in northern Germany identified in the previous phases remain largely in place. The pattern of hot spots does now include several regions in the state of Saxony.

### G Height winter surge 2020-11-14 - 2020-12-23

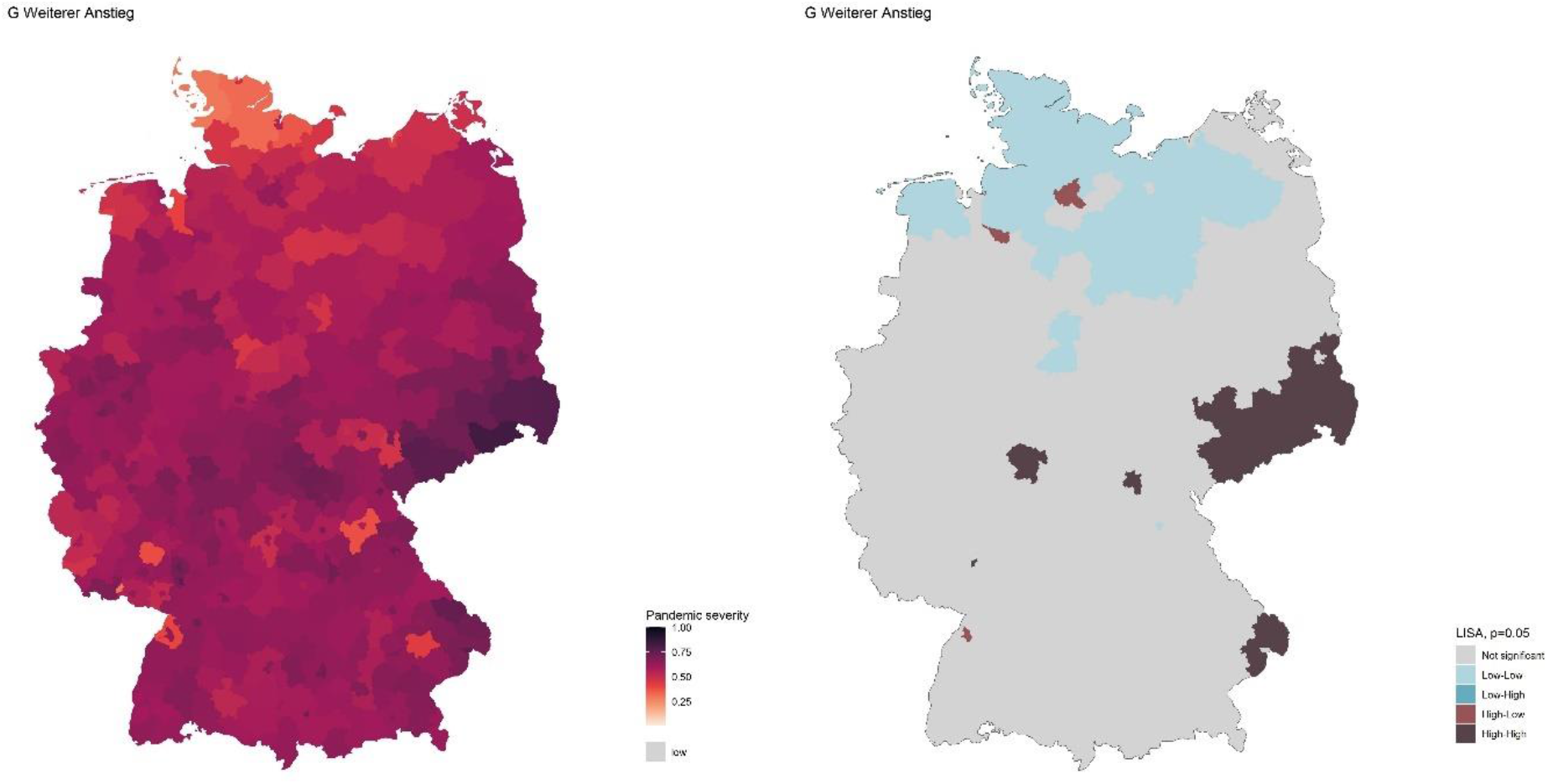

Around mid-November 2020, the autumn surge entered a third phase that lasted till Christmas 2020 and was characterized by a slightly lower growth rate. This phase marks another escalation of the second wave in Germany, as the rapid increase in pandemic severity continues, albeit at a somewhat slower pace. During this phase, variant B.1.1.7 or VOC Alpha is first detected in Germany, albeit without having a marked impact on infection numbers for now. The end of this phase marks the peak of the second wave of COVID-19 in Germany after three months of rapid increase. The rapid increase of pandemic severity was met by more stringent non-pharmaceutical measures, so that the stringency index hit a new peak from early December onwards. This resulted in a decline of average contacts, which, however, remained on substantially higher level compared to the height of the first wave.

This phase is also separated from the previous one by the higher prevalence of outbreaks, notably outbreaks in closed environments such as elderly care facilities, and a high case-fatality rate.

Spatially, this phase is much more concentrated than the previous one, with the spatial autocorrelation of the severity index reaching almost double on average compared to the previous phase. This can be explained by the deteriorating situation in the Saxony cluster as well as some other local hotspots, notably in Lower Bavaria^3^, while the cold-spots large remain in place.

### H Decline winter wave 2020-12-24 - 2021-03-02

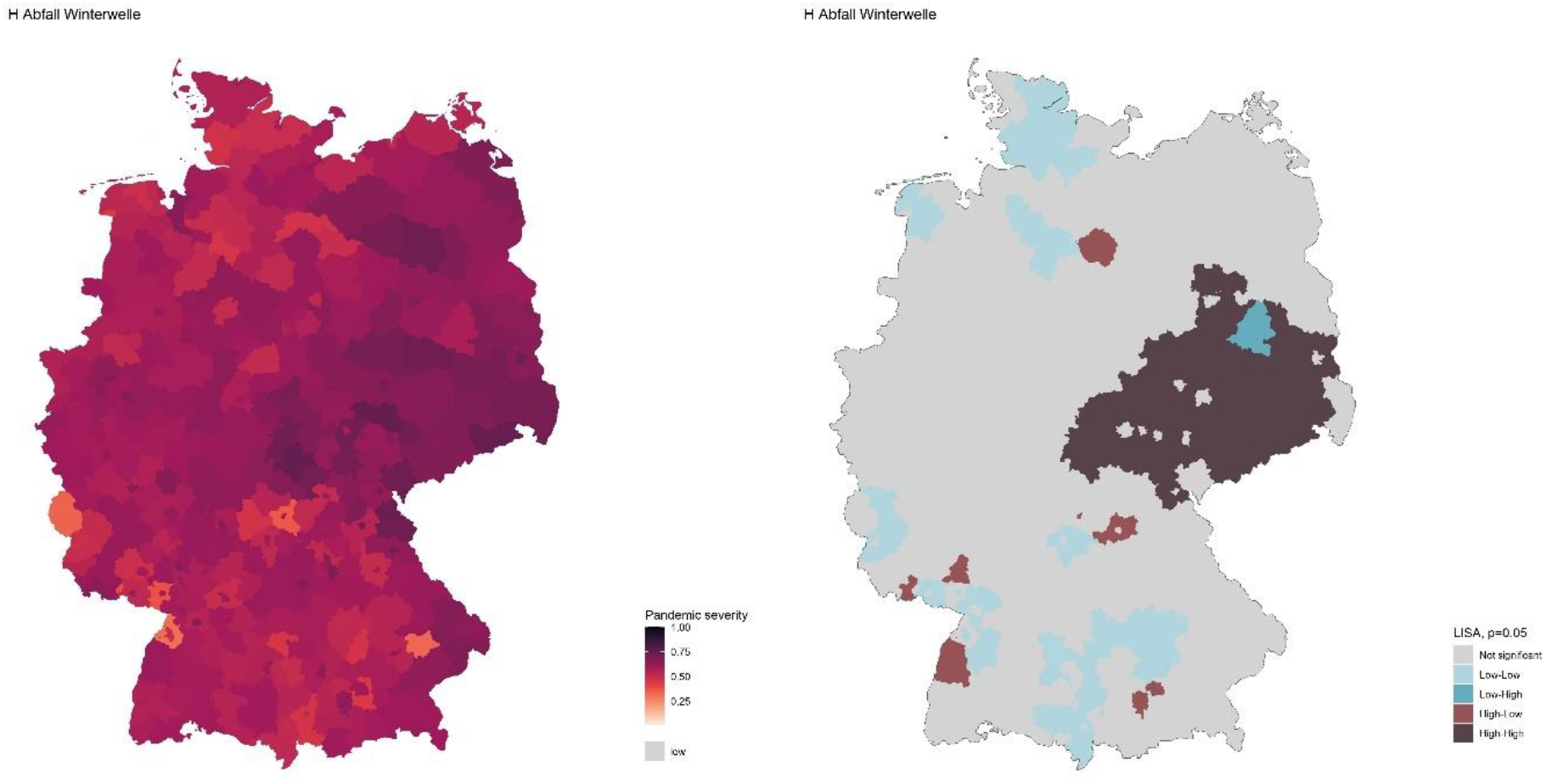

The pandemic severity index hit its second peak between Christmas 2020 and New Year, marking the next phase, which is characterized by decline in pandemic severity over 69 days mostly in January and February 2021. The quick decline of pandemic severity was facilitated by strict non-pharmaceutical measures that were even slightly stricter compared to the previous phase and stayed in place for almost the whole duration. The short time lag between the implementation of strict measures in early December 2020 and the decrease of pandemic severity from mid-December onward implies an effect, although the peak of this wave might be somewhat obscured by the Christmas holidays. Further, the vaccination campaign started in the last days of 2020 and was able wo reach about 4.5 million people in Germany deemed especially vulnerable by the end of this phase. However, the number of outbreaks in closed environments remained high and even increased in January, reaching its peak at 12.6% in the third week of 2021. A high percentage of infections among vulnerable patients in care facilities might also be one of the explanations of a further increase in the case-fatality rate to 3.6%. This value is lower than during the peak of the first wave, however still very high, especially considering the improvements in testing regime in the meantime. Curiously, the overall decline in pandemic severity coincided with an increase in prevalence of the variant B.1.1.7 or VOC Alpha, which already made up about 70% of cases at the end of this phase, after being relatively obscure at the beginning.

Spatially, the hotspot regions in Saxony expanded to the states of Saxony-Anhalt, Brandenburg, and Thuringia, also resulting in a decrease of global autocorrelation, while new cold spots emerged in Southern and Western Germany.

### I Surge Alpha wave 2021-03-03 - 2021-04-28

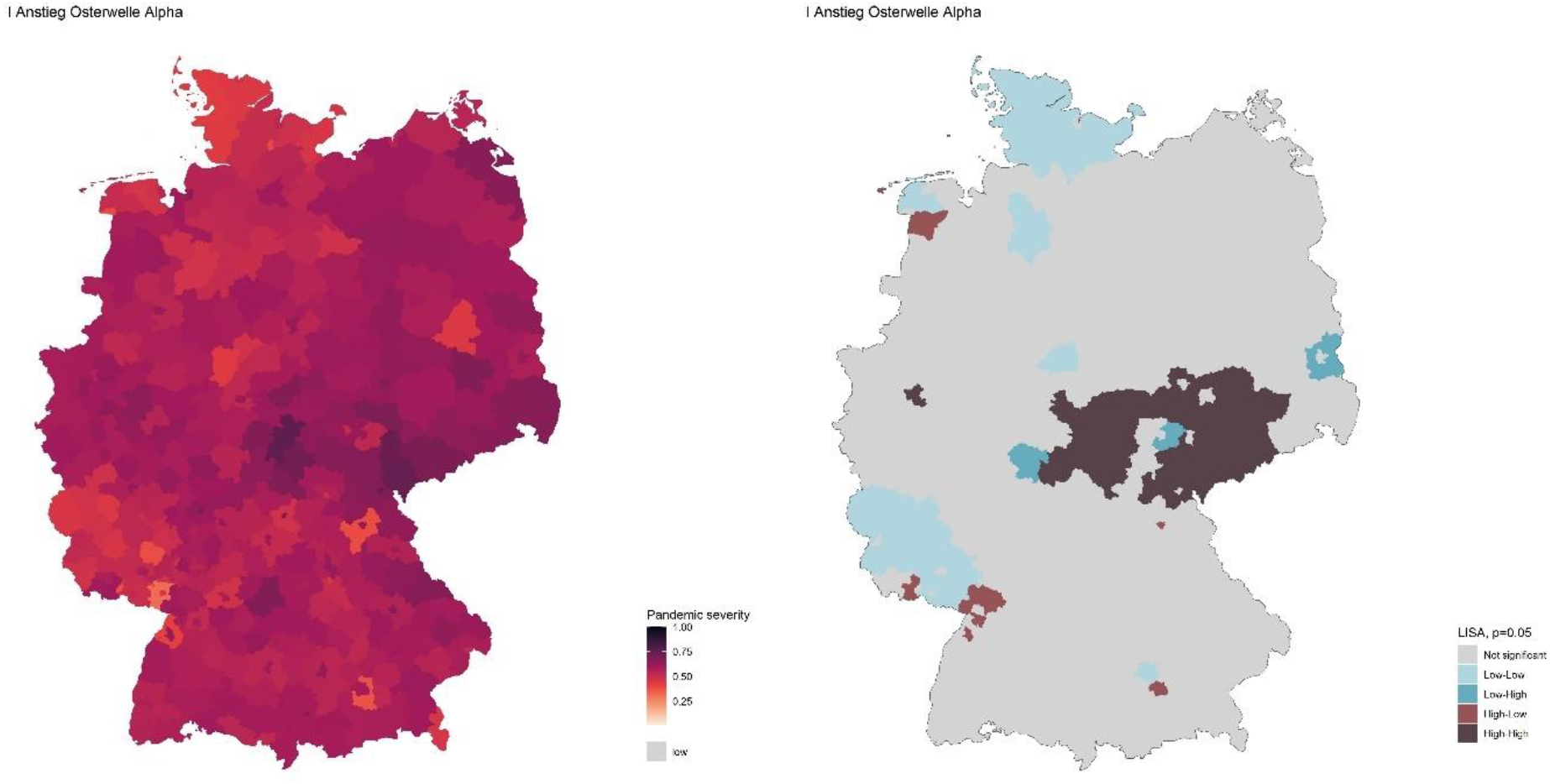

After two months of decline, the pandemic severity index increased again during 57 days in March and April 2021. Since most non-pharmaceutical measures largely staid in place and average social contacts were even lower compared to the previous phases, the increase in pandemic severity can be attributed to the ongoing spread of the variant B.1.1.7 or VOC Alpha. VOC Alpha is more contagious than the clades circulating previously (Campbell et al. 2021) and reached a dominating prevalence during this phase.

The vaccination campaign accelerated during this phase, so that at the end of April around 30 million doses were administered in Germany already. Another important change is the wide-spread implementation of antigen testing in several 1000 state-sponsored testing facilities. These changes and the higher share of vaccinations among the vulnerable population translates into a substantial decrease of the raw case fatality rate, which is more than halved compared to the previous phase. Further, the average increase of pandemic severity during this phase was substantially lower than during previous surges.

The spatial pattern of pandemic severity shifted somewhat compared to the previous phase. While the global spatial autocorrelation decreased slightly, the number of counties identified as hotspot region declined. Most relevant remained the cluster in central Germany comprising of parts of the states of Saxony, Saxony-Anhalt, and Thuringia.

### J Decline Alpha wave 2021-04-29 - 2021-06-16

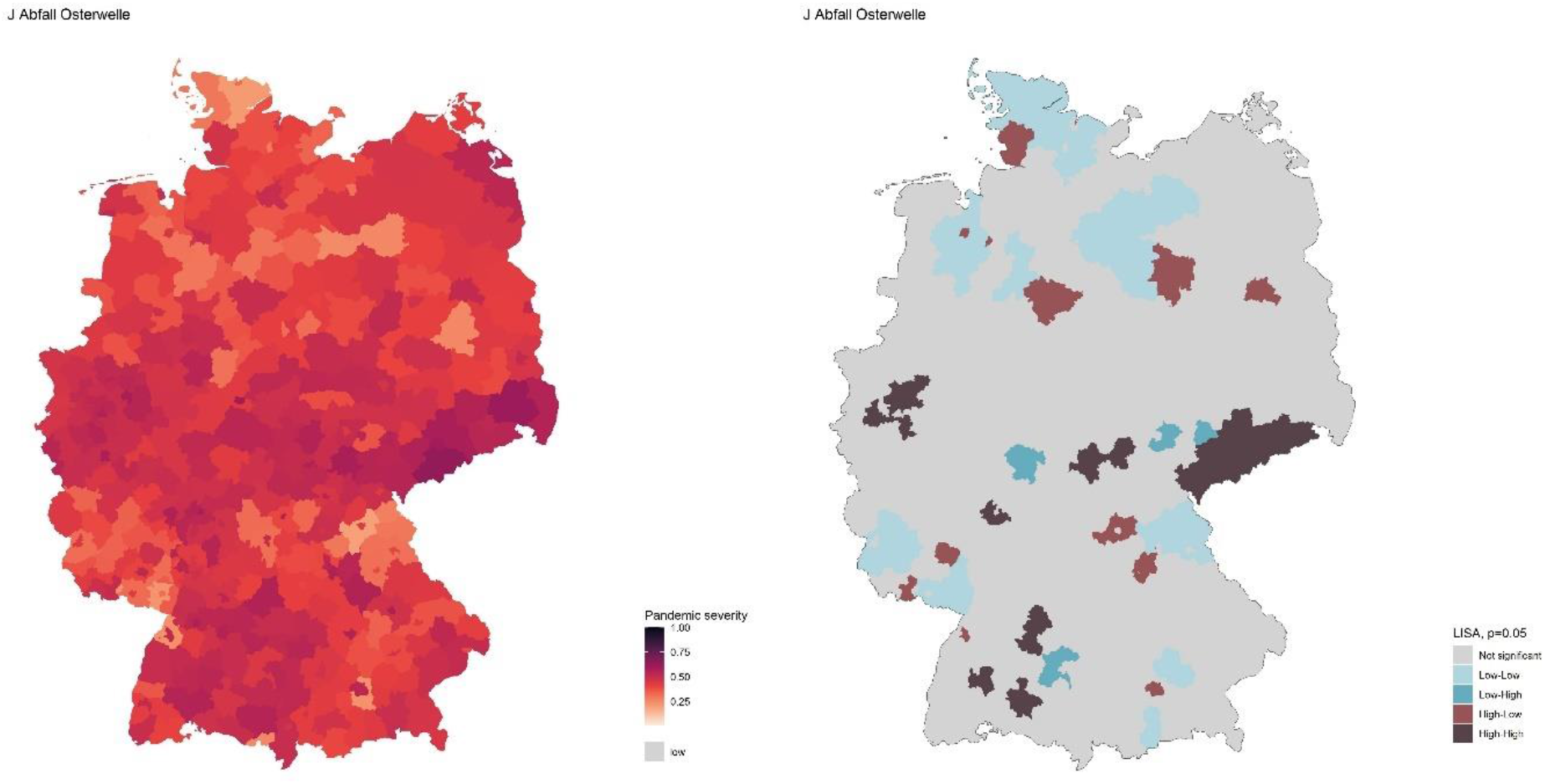

During the second half of April 2021, the third wave of COVID-19 in Germany peaked, inducing again a phase of steep decline in pandemic severity. Compared to the previous phases of decline in pandemic severity, the average decline during the 49 days of this phase was even steeper with -0.008 points per day. While the prevalence of the variant B.1.1.7 or VOC Alpha was dominant throughout the phase, variant B.1.617 or VOC Delta rose in prevalence and was already responsible for about a quarter of infections in mid-June.

The decline in pandemic severity occurred despite a further reduction of non-pharmaceutical measures and an increase of average social contacts. However, the vaccination campaign was further accelerated, so that by the end of this phase about 50% of the German population had received at least a first vaccination. In contrast to the previous phases of steep decline in pandemic severity (phase C and H respectively) this phase was characterized by a further decline in raw case-fatality rate (0.78%), whereas during the first two waves the raw case fatality rate had peaked during the phases of decline. Similarly, the share of cases attributed to outbreaks in closed environments declined further.

The spatial pattern of pandemic severity shifted substantially in this phase. While the spatial autocorrelation declined further slightly, the distribution of hotspot regions changed, as the cluster in central Germany largely dissolved and several dispersed hotspot regions emerged all over Germany.

### K Summer plateau 2021-06-17 - 2021-07-17

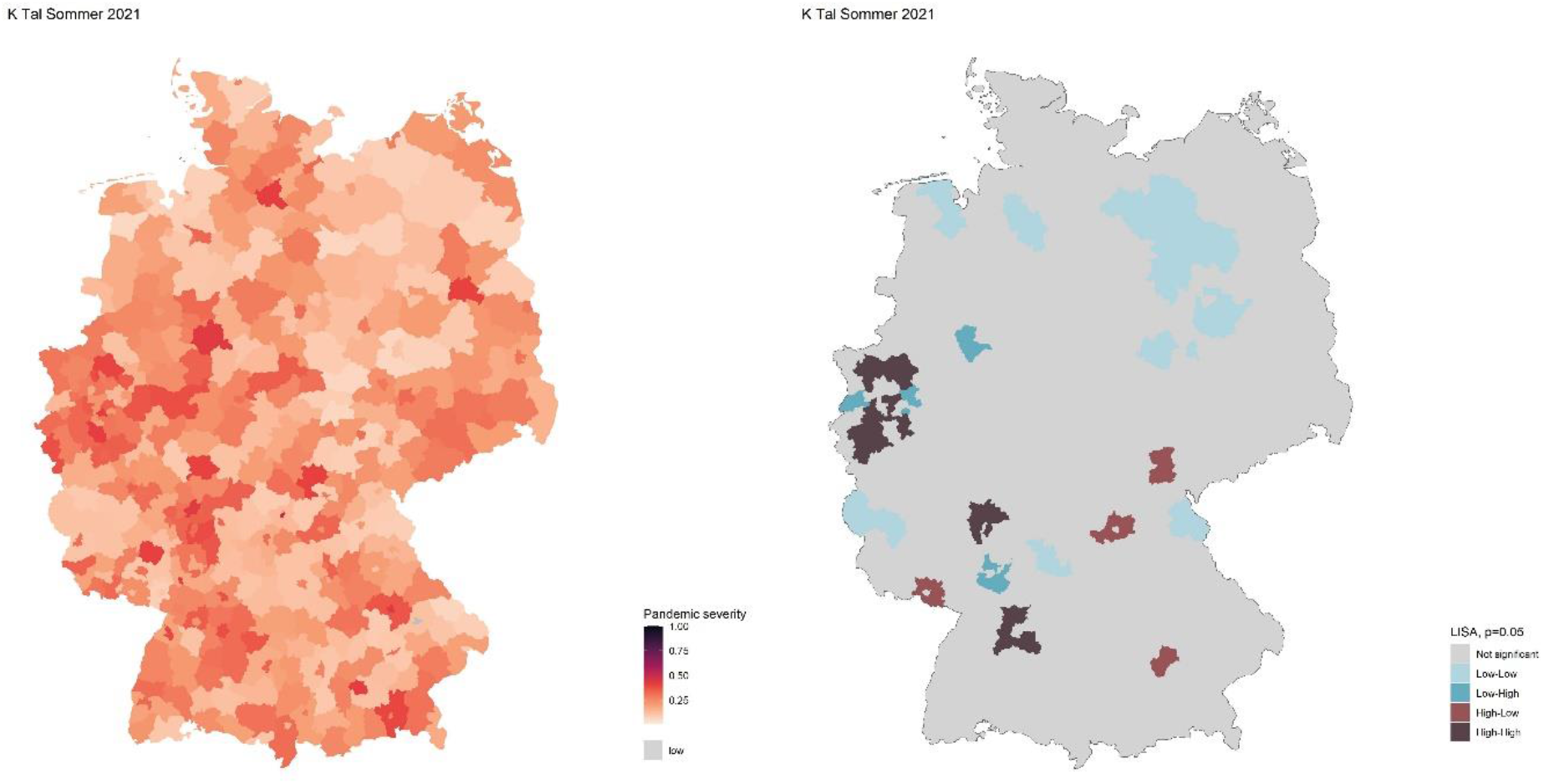

The phase of low and further decreasing pandemic severity during the summer of 2021 was shorter compared to 2020 and lasted only 31 days between mid-June and mid-July. During this phase the variant B.1.617 or VOC Delta reached dominating prevalence, setting the preconditions of subsequently rising pandemic severity.

At the same time, the vaccination campaign further accelerated, so that in July about two thirds of the population had received at least a first dose of vaccine and about half were fully vaccinated. Non-pharmaceutical measures were further reduced, albeit not as much as in the previous summer. However, average social contacts increased only slightly, remaining well below the levels of the previous summer let alone pre-pandemic levels.

This phase is characterized by a drastic decrease in spatial autocorrelation, to the lowest level among all fifteen phases. As in the summer of 2020 dispersed outbreaks prevail. Notably, the East German cluster in Saxony is no longer significant in our data.

### L End summer plateau 2021-07-18 - 2021-08-10

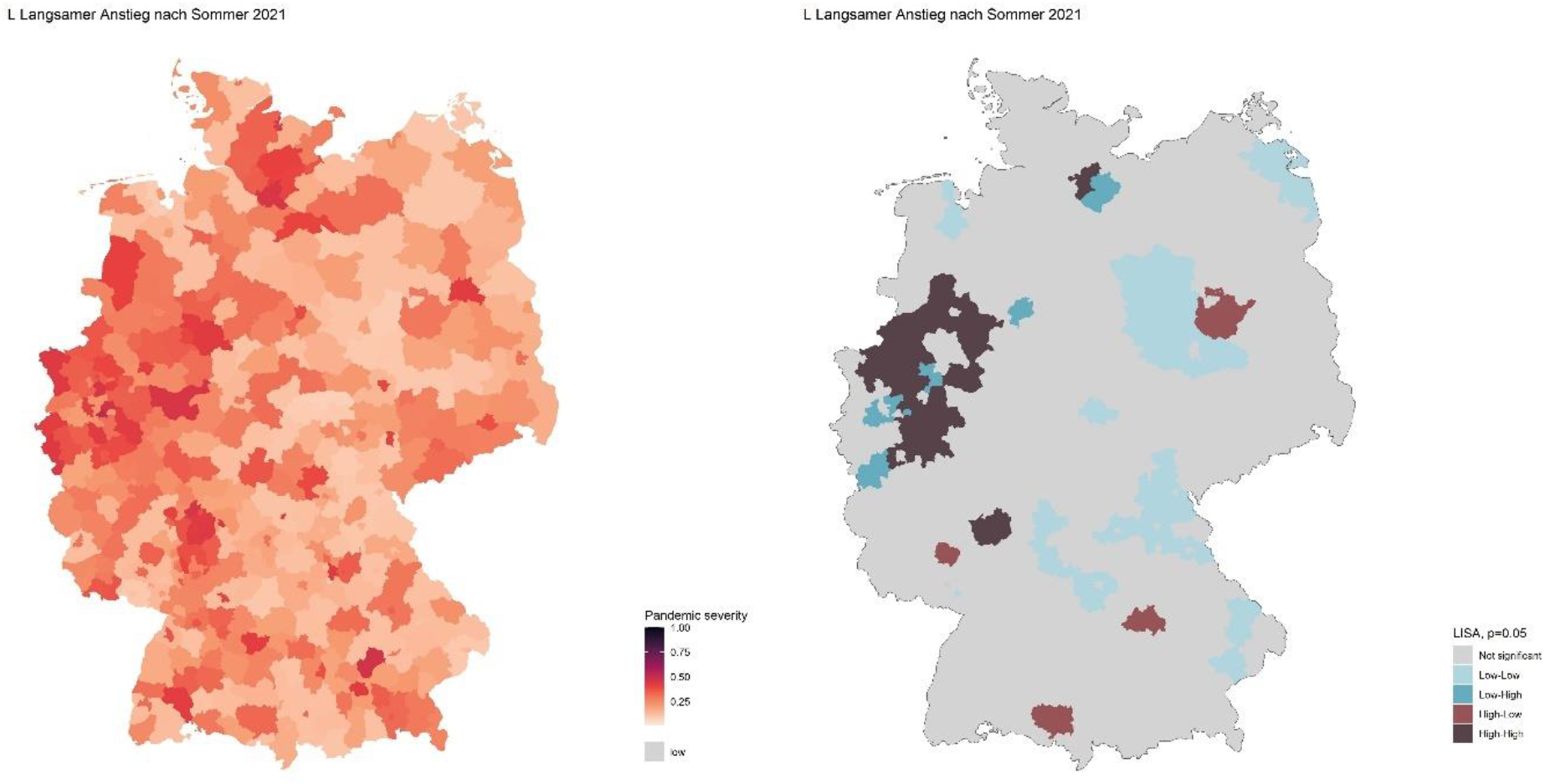

The second phase during the summer of 2021 is characterized by a slow increase in pandemic severity. After the decreasing trend reversed around mid-July, the increasing trend does not accelerate for another three weeks. Variant B.1.617 or VOC Delta continues to gain ground and is responsible for about 99% of infections by mid-August, which is the most homogeneous distribution of variants in Germany up to that point. While the average raw case-fatality rate during this phase further decreased and is the lowest among all phases with 0.42%, most other conditions remained unchanged from the previous phase.

The spatial autocorrelation increased but remained comparably low. Most dispersed hotspots changed compared to the previous phase. However, some hotspots in North Rhine-Westphalia persisted and a regional cluster emerged.

### M Entry Delta wave 2021-08-11 - 2021-10-24

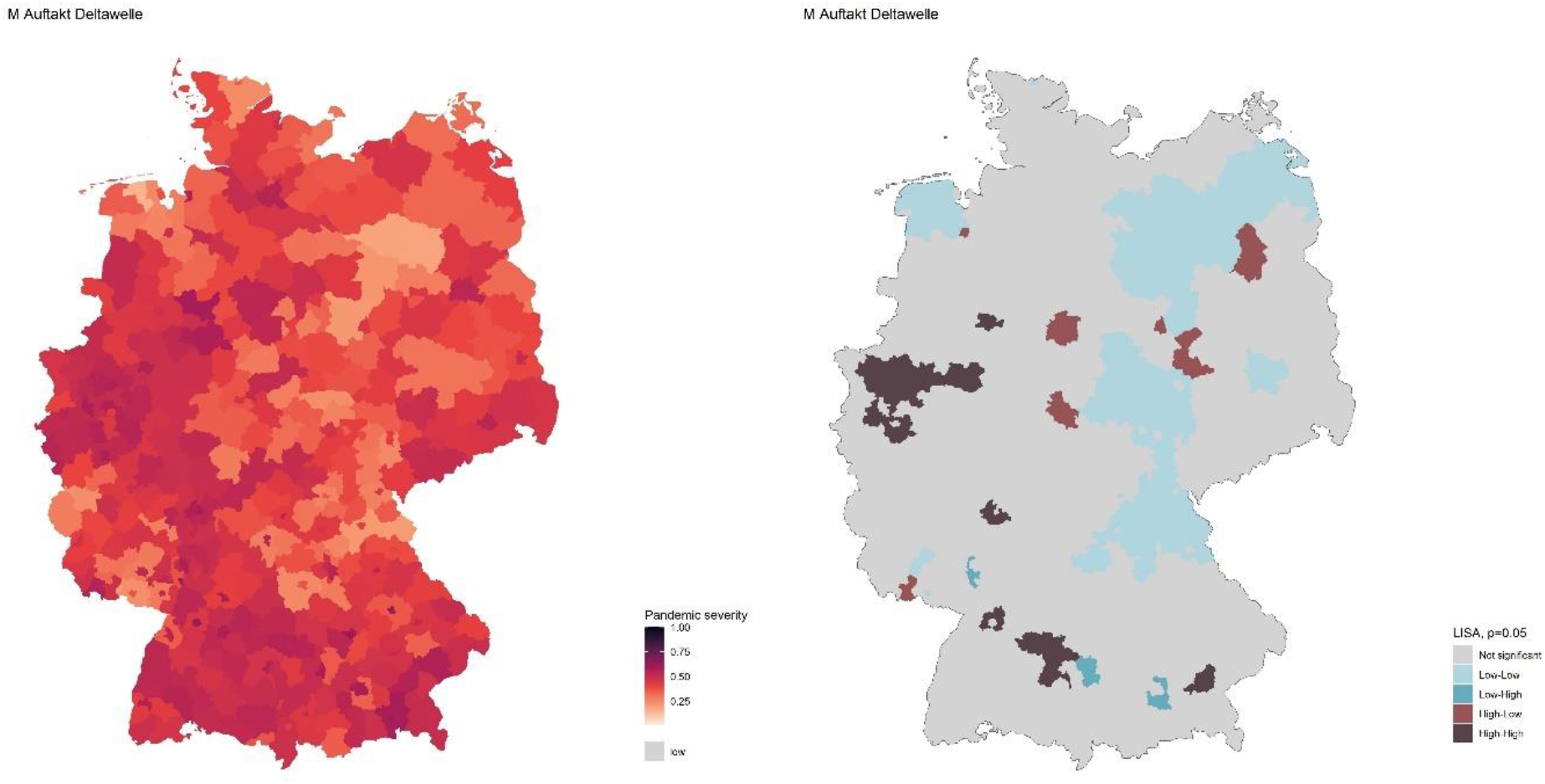

In August 2021, the pandemic severity index began to rise faster than in the previous phase. However, the entry phase to the coming delta-wave is more complex, since it is characterised by two distinct periods of rapid increase in pandemic severity separated by three weeks of stagnation and even decline during September, so that the average increase over the 75 days is relatively low. Variant B.1.617 or VOC Delta continues to dominate, being responsible for virtually all infections during this phase.

Despite the increases in pandemic severity, the level of non-pharmaceutical measures was lowered continuously, until it reached its lowest level since the beginning of the pandemic in early October 2021. However, average social contacts did not increase accordingly. The vaccination campaign decelerated substantially during this phase.

While the spatial autocorrelation somewhat increased, this phase is again characterized by dispersed hotspots and a cluster of hotspot regions in North-Rhine Westphalia. Northern and central parts of Germany show cold spots.

### N Surge Delta wave 2021-10-25 - 2021-12-02

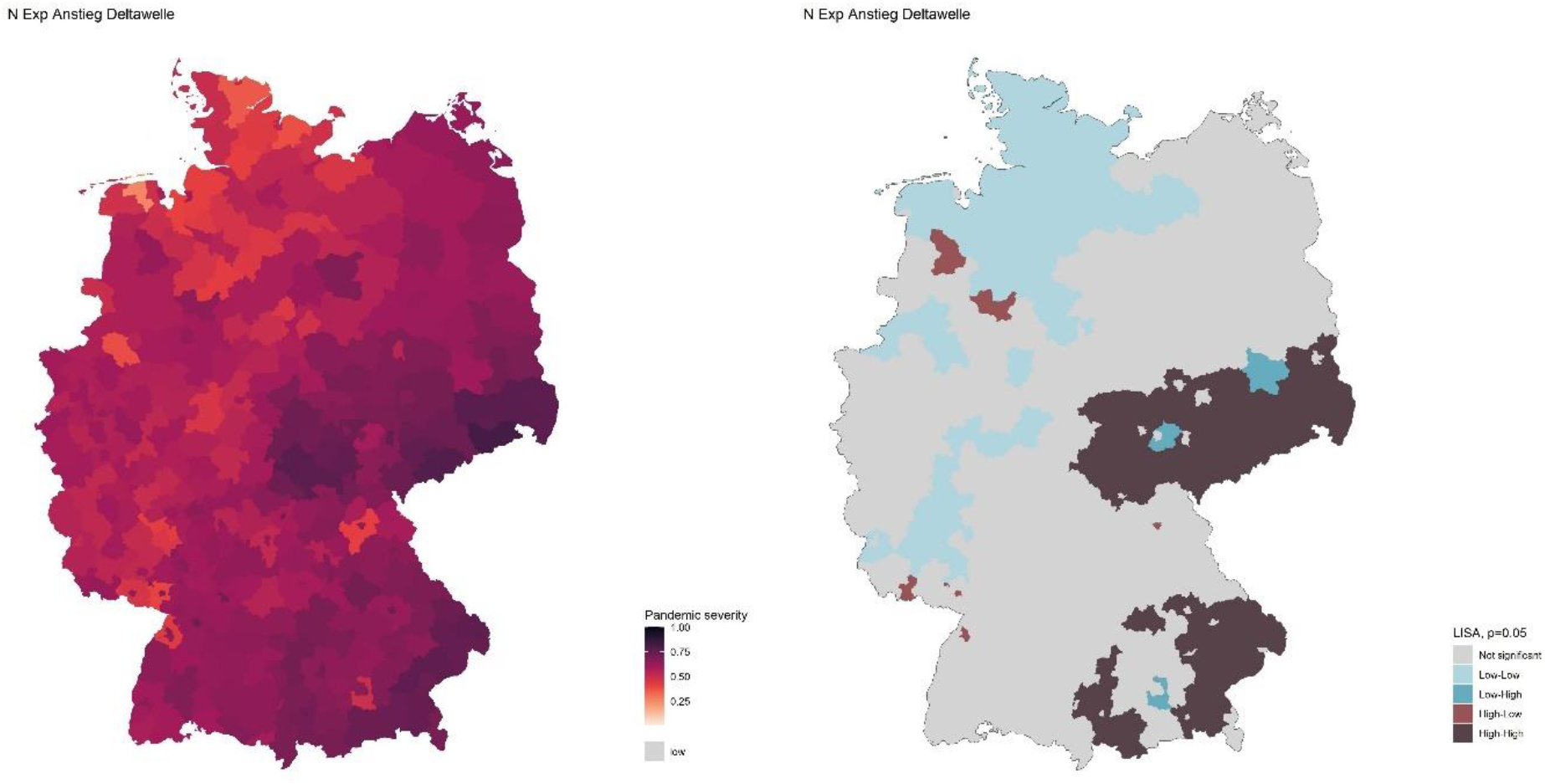

During the second half of October and all of November of 2021 another rapid surge in pandemic severity occurred. With an average daily increase of 0.013 points per day over 39 days, this marks both the steepest increase in pandemic severity and the highest overall pandemic severity throughout the years of 2020 and 2021. Variant B.1.617 or VOC Delta continues to dominate, although the first cases of variant B.1.1.529 or VOC Omicron were observed in Germany in late November. As during the previous surges, the raw case-fatality rate increased. However, it remained substantially below the level during other surges with a value of 0.96.

In reaction to the rapid surge in pandemic severity, the level of non-pharmaceutical measures was increased again, albeit peaked well below the levels seen in the previous waves. Similarly, the vaccination campaign that had somewhat stalled in the previous phase was accelerated by distributing booster vaccinations, while the prevalence of vaccinations did remain around 75% of the population in Germany.

Remarkably, the cluster of hotspot regions in central Germany reappeared during this phase together with another large cluster of hotspot regions in Southern Bavaria. reappearance can be localized in Bavaria around the area of Passau. In the northern part of Germany cold spots prevail, especially in the states of Schleswig-Holstein and Lower Saxony. These trends explain a drastic spike in global spatial autocorrelation to the highest value among all phases.

### O Decline Delta wave 2021-12-03 - 2021-12-30

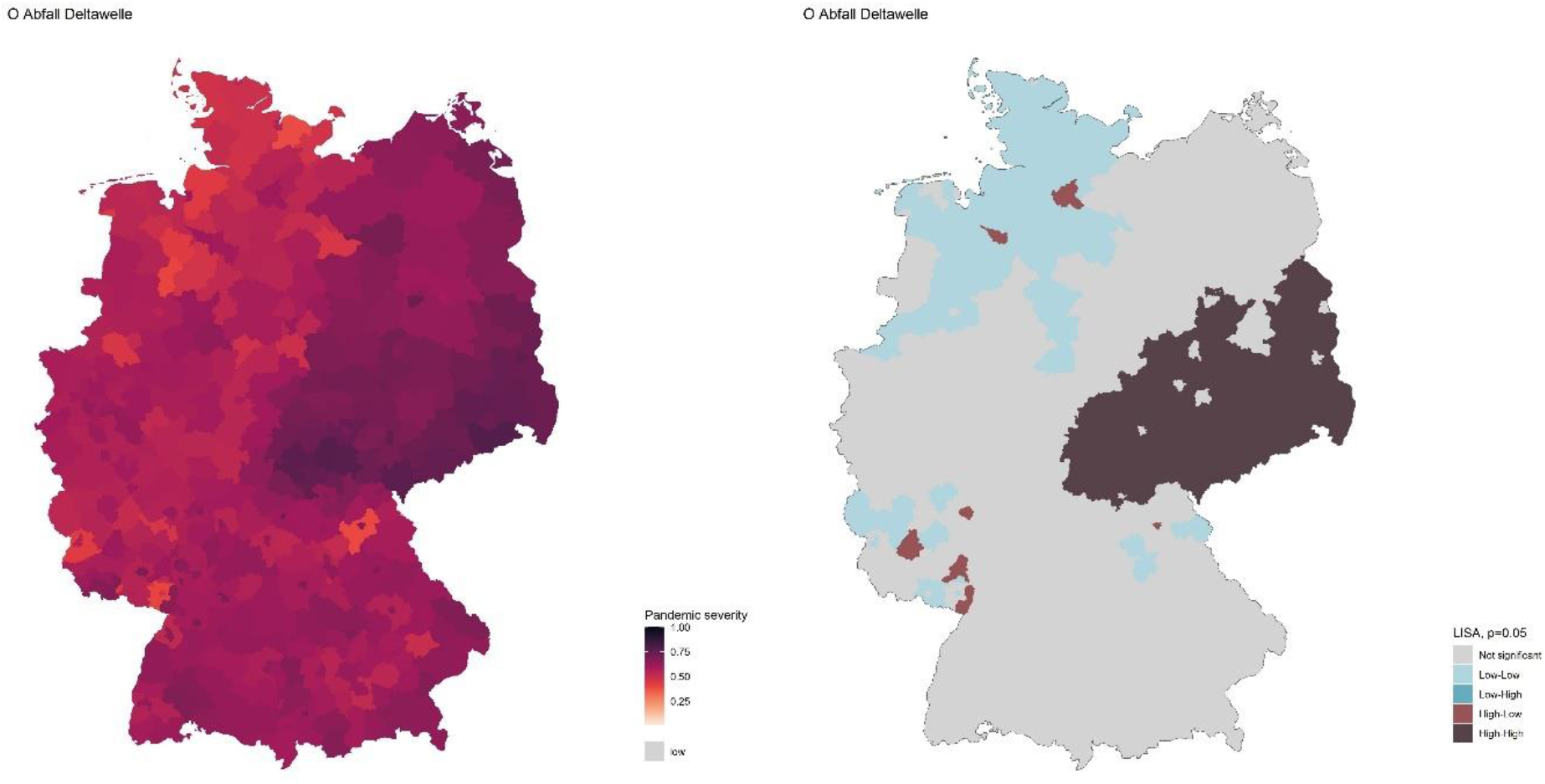

The final phase of the COVID-19 pandemic in 2021 marks the decline of the delta wave in December 2021. Like the rapid surge in the month before, the decrease of pandemic severity during the 28 days of this phase was the fastest on average among all phases. While variant B.1.617 or VOC Delta still dominated this phase, variant B 1.1.529 or VOC Omicron gained traction quickly and already had a prevalence of 41% of the cases at the end of December.

The level of non-pharmaceutical measures remained on the level reached during the previous phase and the index of average social contacts increased only slightly. The vaccination campaign proceeded by adding a substantial amount of booster vaccinations, so that at the end of 2021 almost 45% of the population had reached triple vaccination status.

While the cluster of hotspot regions in southern Bavaria has disappeared in this phase, the cluster of hotspot regions in central Germany has enlarged to the north and the west. The cold spots in northern Germany largely remain in place, although the cities of Bremen and Hamburg emerge as dispersed hotspots, likely due to early outbreaks of VOC Omicron. Due to these spatial patterns, the global autocorrelation remains on a very high level.

## Discussion and conclusion

This working paper presents a phase model describing the COVID-19 pandemic in Germany during the years 2020 and 2021 in order to analyze the tempo-spatial dynamics of the pandemic. Based on an index of pandemic severity, our analysis identified fifteen distinct pandemic phases, which were subsequently analyzed by using additional data on dominant clades of SARS-CoV-2, outbreaks, mobility, and countermeasures. In this section, the suitability of the pandemic severity index is evaluated before the findings on tempo-spatial variation of the COVID-19 during our study period are summarized and discussed from a socio-spatial process perspective.

Initially, the pandemic severity index was introduced to add robustness to the pandemic modelling. In comparison to models using infection incidence data only, the pandemic severity index allows for better comparability in time due to changed conditions. For example, changes in the testing regime or can lead to drastic biases in case numbers: While the data on the first wave implies a very low incidence of COVID-19 cases, the numbers of deaths and intensive-care patients are much higher in relation, so that our composite index indicates a higher pandemic severity compared to the case numbers. In the aftermath of pandemic waves, the pandemic severity index declines slower compared to case numbers, which reflects longer stays in intensive care and delayed deaths. Therefore, our severity indicator provides a more robust representation of the overall pandemic activity and enables to compare pandemic dynamics in time and space.

Considering the spatial patterns between the different phases, our analysis unveiled an unequal distributed of pandemic activity over time and space. Spatial autocorrelation is generally higher during the phases of surging pandemic severity. Every surge had its significant hotspots and cold spots, only some of which were consistent over time. Especially the first wave had a relatively distinct spatial pattern of dispersed hotspots in southern and western Germany. In contrast, the three following pandemic waves displayed some similarities in terms of spatial patterns. During almost all surges of these waves a relatively coherent cluster of hotspot regions was found in central Germany (including the states of Saxony, Thuringia, Saxony-Anhalt, and Brandenburg). Similarly, relatively persistent cold spots were found in northern Germany (including the states of Mecklenburg-Western Pomerania, Schleswig Holstein, and Lower Saxony). Curiously, both the regions with dominant hotspots and cold spots were relatively stationary (see also Kuebart & Stabler 2023). While the approach to analyze tempo-spatial patterns based on the local indicators of spatial association of phases of pandemic activity generally worked well, the LISA analysis was somewhat distorted by small urban counties (Kreisfreie Städte), which small size and geometry (most of them have only one neighboring county). Nevertheless, an analysis of spatial patterns on the basis of NUTS 2 areas or other regions of higher spatial order (e.g., Raumordnungskategorien) might be more suitable.

Considering the results from a socio-spatial perspective, four sub-processes can be seen as driving forces for the tempo-spatial dynamics of the pandemic. First, the process of infecting is obviously at the core of each pandemic. Infecting is bound on several requirements, including a tie between two or more persons and suitable conditions at the place of co-location (e.g., temperature, humidity, ventilation). What constitutes a tie and which conditions are suitable for infections depends on the characteristics of the pathogen, often subsumed under its transmissibility. In the case of SARS-CoV-2, the co-location of persons was necessary for infections to occur. However, these characteristics were not stable over the course of the pandemic, since the specific characteristics of the pathogen itself changed with evolutionary pressure, so that new clades of SARS-CoV-2 emerged (Eales 2022), which differed in transmissibility characteristics (Campbell et al. 2021).

Second, the process of networking influences how pathogens diffuse since the ties necessary for infections are determined by the existing ties within societies (Bian 2004). COVID-19 has been found to have an extremely positively skewed distribution of out-degree nodes (Jo et al. 2021). The large prevalence of contact in trans-local networks facilitate the rapid movements of new pathogens (Kuebart& Stabler 2020A). Since transmission not only depends on the existence of ties but also the quality/intensity of ties (Lloyd-Smith et al. 2005). For example, a variant of SARS-CoV-2 with a higher transmissibility can spread between two individuals that only briefly interacted, whereas less transmissible variants required longer or more intense interactions (Lindstrøm et al. 2021; Campbell et al. 2021) Therefore, the shifting characteristic of the pathogen directly affects the network space, in which the pathogen spreads and have to be considered for a tempo-spatial analysis.

Third, the specific characteristics of places impacts local infection dynamics. Place-base scaling can facilitate rapid increase of pandemic severity in a region when a large group of persons gathers at a place with favorable conditions for infection (Kuebart&Stabler 2020B). Such super-spreading events were frequently observed throughout all phases of the pandemic in settings such as night-clubs, fairs, or churches. Notably, super-spreading was not bound to phases of high pandemic severity, as a single infectious person is sufficient to trigger multiple infections when certain conditions are met. On the other hand, place-based networking occurs when a network is bound to a certain place, such as a ship or an elderly care facility (Kuebart& Stabler 2020A). Here, the place does not necessarily facilitate a large number of infections at the same time, but rather complicates the protection of individuals once the pathogen has entered the network. Such outbreaks in closed environments were found to rise in frequency with a certain time lag after pandemic severity increases. While relatively few cases were attributed to outbreaks in closed environments, a high number of deaths can be attributed to them, due to the higher individual risk for persons in elderly care facilities. This effect seems to subside with the third wave, likely due to progress in the vaccination campaign.

Fourth, the societal responses to the pandemic shaped the spread of COVID-19 substantially through territorialization. These responses were visible both in form of specific policies on non-pharmaceutical interventions implemented by national or regional governments (e.g., social distancing rules) but also in form behavioral changes in the population to limit the spread of the disease (e.g., caution in contact with vulnerable individuals). Of course, each specific policy had a behavioral change in mind, so that compliance to the policies is another important dimension to this factor. From a spatial perspective, both the policies set and the compliance with them are bound to specific territories and the implementation of policies regarding COVID-19 depended on socio-political conditions (Ren 2020). While it would be misleading to put too much emphasis on the territorial dimension of space in the era of “post-Westphalian pathogens” (Fidler 2003), the “societal immune system” visible during the COVID-19 pandemic has proven the relevance of territories and their impact on the pandemic process (Kuebart & Stabler 2021).

In conclusion, both spatial impact factors such as outbreaks or non-pharmaceutical interventions and their compliance and temporal effects such as subsequent waves have influenced the dynamics of the COVID-19 in Germany. Therefore, a tempo-spatial perspective that conceptualizes the pandemic as a socio-spatial process seems to be a promising approach to gain insights into how, when and where the pandemic unfolds. Further research might especially benefit from better availability of data, since especially data on outbreaks and imported infections is only available very limited at the time of publication. In this context, especially the limited willingness of federal authorities to provide existing dataset was unhelpful. Another limitation of this study is the scale of analysis. Both more fine-grained analysis of hotspot regions and analysis on a higher spatial order is necessary to better understand the dynamics of the pandemic. Finally, further development of the pandemic severity indices to incorporate more variables or other sub-indices seems to be a path for further research into pandemic geographies.

## Data Availability

All data produced in the present study are available upon reasonable request to the authors. All raw data are part of the public domain already.

## Footnotes

1 https://www.corona-datenplattform.de

2 Hodgecroft (2021), all variants are described using the using Pango nomenclature system

3 Notably a cluster in the Passau region: m.buergerblick.de/nachrichten/landkreis-passau-zehn-corona-tote-in-zehn-tagen-a-64151.html

